# Factors Associated with COVID-19 Mitigation Behavior among US Adults

**DOI:** 10.1101/2020.07.20.20157925

**Authors:** Debra C. Lemke, Klaus W. Lemke

## Abstract

In early 2020, the CDC issued guidelines for personal COVID-19 mitigation behavior, such as mask-wearing, hand-washing, and social-distancing. We examine individual socio-behavioral factors that potentially predict mitigation compliance using public data. Our analysis finds that pandemic prompted strong mitigation behavior by adults, especially among females, non-whites, urban dwellers, and the psychological unwell. Other positive predictors were post-secondary education and higher income. Health symptoms and clinical risk factors did not predict increased mitigation practices, nor did age 65+ and proximity to infected persons. Our study findings are congruent with a report that pointed to a lack of increased pandemic mitigation practices in households with confirmed infections and health risks. We also point to lower levels of psychological resilience, lower socio-economic status, and non-urban location as potential explanatory factors for lack of mitigation behavior. Understanding what factors are associated with mitigation behavior will be important for policy makers in their efforts to curb the COVID-19 pandemic.

## Background

In January 2020, the US declared the coronavirus outbreak a public health emergency and subsequently the CDC issued guidelines for personal mitigation behavior, such as mask-wearing, hand-washing, and social-distancing.

## Objective

We examine individual socio-economic factors that potentially predict mitigation compliance using public data.^1^ We hypothesize that health risk factors, presence of symptoms, and psychological wellbeing predict mitigation behavior. Understanding what factors are associated with mitigation behavior will be important for policy makers in their efforts to curb the COVID-19 pandemic.

## Methods and Findings

We used public data from the COVID Impact Survey (CIS), conducted by NORC at the University of Chicago for the Data Foundation.^1^ Several CIS questions were taken from prior national surveys including the American Community Survey, Behavioral Risk Factor Survey, and Current Population Survey.^2^ CIS data for samples of adults aged 18 and older were collected between late April and early June 2020. NORC provided sampling weights.

We combined questionnaire items into a set of parsimonious summary scales and scores. A *mitigation compliance scale* summed 17 behavior items; several were used previously by the CDC for the 2009 National H1N1 Flu survey. Our summary *symptoms score* used 8 self-reported symptoms: fever, chills, shortness of breath, cough, sore throat, fatigue or tiredness, muscle pain, and loss of taste and smell. These symptoms were widely recognized indicators for potential SARS-CoV-2 infection. The CDC further identified risk factors for severe COVID-19 illness.^3^ Our *risk factor score* counted self-reported diabetes, high blood pressure or hypertension, heart disease or stroke, asthma, chronic lung disease or COPD, bronchitis or emphysema, liver disease, cancer, and immunocompromised condition. Lastly, we aggregated CIS items on experiencing feelings of depression, anxiety, hopelessness, loneliness, and stress due to the coronavirus crisis within the last 7 days into an *anxiety-depression-stress scale* for this study.

Our objective was to develop models to predict mitigation behavior. A base model regressed the mitigation compliance scale on symptom and risk factor scores. Successive models added independent variables including the anxiety-depression-stress scale and dichotomous variables for age 65 and older, individual characteristics (female gender, non-white race, post-secondary degree), household income greater than $50,000, urban location, and household member has COVID-19. We performed our analyses in SPSS version 26.

Nearly all adults (99%) in the survey reported mitigation behavior. The median number of protective practices was 8 (range=17). The mitigation compliance scale reliability was greater than 0.72 and increased to 0.80 in wave 3. Respondents reported 1.0-1.2 (SD=1.4) symptoms on average, and 49-52% were symptomatic. Nearly half (45-49%) also reported at least 1 risk factor (range=9) consistent with BRFSS estimates.^4^ CIS percentages for cancer, chronic lung disease, diabetes, heart disease, hypertension, and liver disease were within 3 percentage points of BRFSS benchmarks; prevalence of asthma, bronchitis or emphysema, and immunocompromised condition were higher in the CIS. Percentages of depression, anxiety, feelings of hopeless, and loneliness were greater than 35% in each wave. The anxiety-depression-stress scale (median=7; range=15) reliability was 0.81-0.86 (Table 1).

**Table 1.** Descriptive Statistic for Mitigation Compliance and Anxiety-Depression-Stress Scales and Computed Risk Factors and Symptom Scores based on the COVID Impact Survey (CIS) in 2020

|  | **CIS Wave 1:**<br>**April 20-26, 2020** | **CIS Wave 2:**<br>**May 4-10, 2020** | **CIS Wave 3:**<br>**May 30- June 8, 2020** |
| --- | --- | --- | --- |
| **Mitigation Compliance Scale** |  |  |  |
| Reliability Coefficient | 0.724 | 0.726 | 0.803 |
| Mean | 8.20 | 7.96 | 7.73 |
| Std. Deviation | 3.26 | 3.27 | 3.48 |
| Median | 8 | 8 | 8 |
| Range | 0 – 17 | 0 – 17 | 0 – 17 |
| Low (0 to 6) | 28.9% | 31.3% | 33.3% |
| Moderate (7 to 9) | 35.8% | 34.5% | 35.2% |
| High (10 to 17) | 35.3% | 34.2% | 31.5% |
| **Anxiety-Depression-Stress Scale** |  |  |  |
| Reliability Coefficient | 0.821 | 0.815 | 0.857 |
| Mean | 7.80 | 7.44 | 7.71 |
| Std. Deviation | 3.41 | 3.08 | 3.45 |
| Median | 7 | 6 | 6 |
| Range | 5 – 20 | 5 – 20 | 5 – 20 |
| Low (5) | 36.6% | 39.5% | 39.0% |
| Moderate (6 to 8) | 33.6% | 33.9% | 32.9% |
| High (9 to 20) | 29.8% | 26.6% | 28.1% |
| **Risk Factors Score** |  |  |  |
| Mean | 0.98 | 0.99 | 0.90 |
| Std. Deviation | 1.24 | 1.18 | 1.21 |
| Median | 1 | 1 | 1 |
| Range | 0 – 9 | 0 – 9 | 0 – 9 |
| No risk factors | 52.4% | 50.5% | 54.8% |
| 1 or more risk factors | 47.6% | 49.5% | 45.2% |
| **Symptoms Score** |  |  |  |
| Mean | 1.16 | 1.07 | 1.01 |
| Std. Deviation | 1.41 | 1.38 | 1.30 |
| Median | 1 | 1 | 1 |
| Range | 0 – 8 | 0 – 8 | 0 – 8 |
| No symptoms | 47.9% | 50.9% | 51.5% |
| 1 or 2 symptoms | 36.9% | 35.5% | 36.6% |
| 3 or more symptoms | 15.2% | 13.6% | 11.9% |

Regression results for models to predict mitigation behavior are shown in Table 2. Greater anxiety-depression-stress, female gender, post-secondary degree, household income above $50,000, and urban location were consistently predictive of more mitigation practices; however, symptoms, risk factors, and age 65+ were not significant predictors across CIS waves. Having a household member with COVID-19 diagnosis was predictive of *fewer* mitigation practices in wave 3.

**Table 2.**
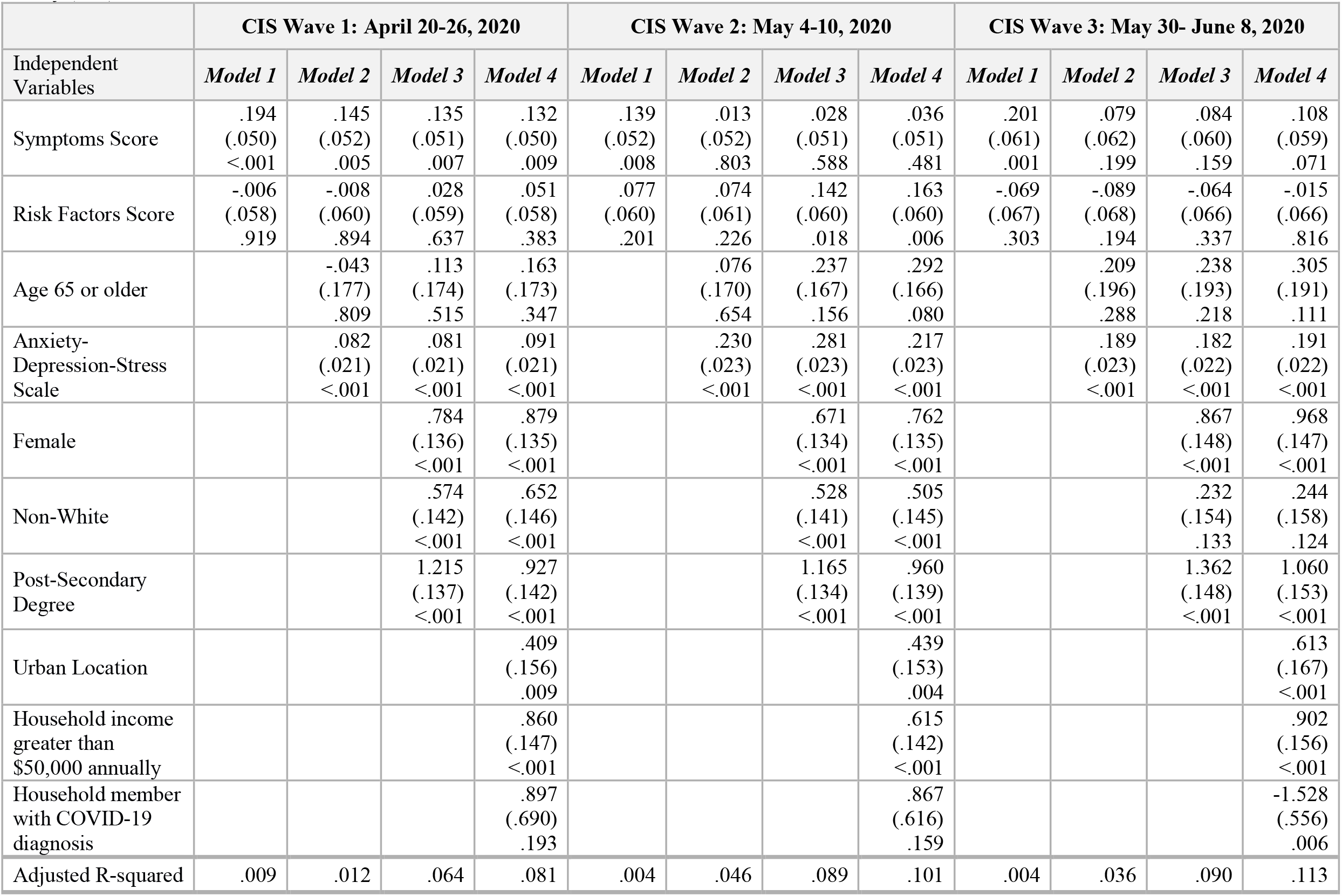
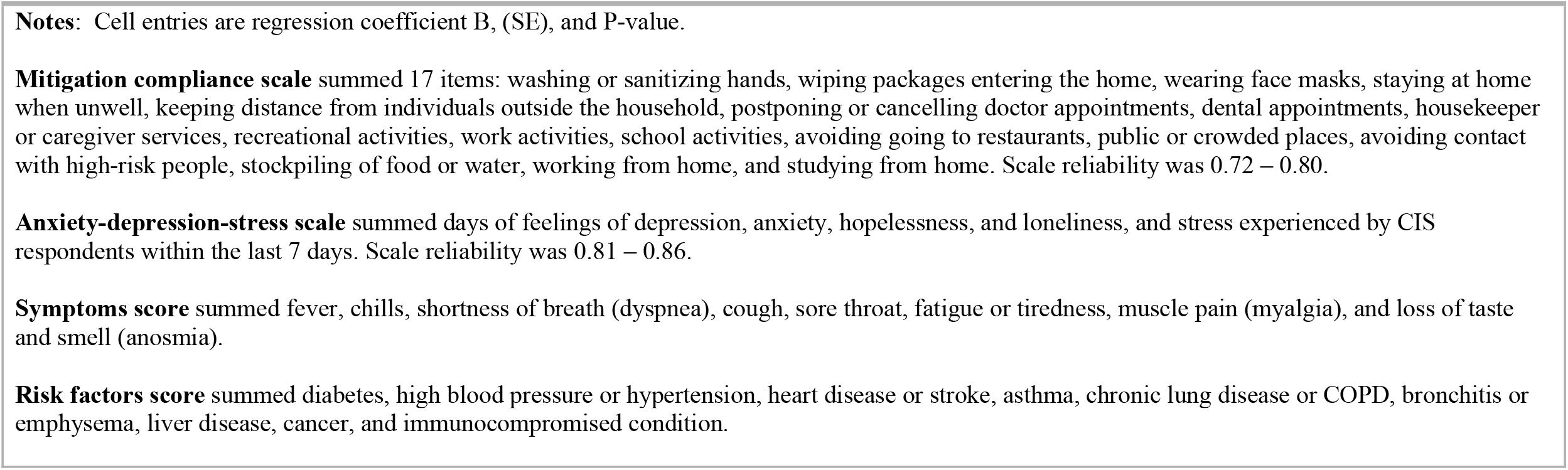
Multivariate Regressions with **Mitigation Compliance Scale** as the Dependent Variable and 4 Sets of Independent Variables, COVID Impact Survey (CIS) 2020.

|  | CIS Wave 1: April 20-26, 2020 |  |  |  | CIS Wave 2: May 4-10, 2020 |  |  |  | CIS Wave 3: May 30- June 8, 2020 |  |  |  |
| --- | --- | --- | --- | --- | --- | --- | --- | --- | --- | --- | --- | --- |
| Independent Variables | *Model 1* | *Model 2* | *Model 3* | *Model 4* | *Model 1* | *Model 2* | *Model 3* | *Model 4* | *Model 1* | *Model 2* | *Model 3* | *Model 4* |
| Symptoms Score | .194<br>(.050)<br><.001 | .145<br>(.052)<br>.005 | .135<br>(.051)<br>.007 | .132<br>(.050)<br>.009 | .139<br>(.052)<br>.008 | .013<br>(.052)<br>.803 | .028<br>(.051)<br>.588 | .036<br>(.051)<br>.481 | .201<br>(.061)<br>.001 | .079<br>(.062)<br>.199 | .084<br>(.060)<br>.159 | .108<br>(.059)<br>.071 |
| Risk Factors Score | -.006<br>(.058)<br>.919 | -.008<br>(.060)<br>.894 | .028<br>(.059)<br>.637 | .051<br>(.058)<br>.383 | .077<br>(.060)<br>.201 | .074<br>(.061)<br>.226 | .142<br>(.060)<br>.018 | .163<br>(.060)<br>.006 | -.069<br>(.067)<br>.303 | -.089<br>(.068)<br>.194 | -.064<br>(.066)<br>.337 | -.015<br>(.066)<br>.816 |
| Age 65 or older |  | -.043<br>(.177)<br>.809 | .113<br>(.174)<br>.515 | .163<br>(.173)<br>.347 |  | .076<br>(.170)<br>.654 | .237<br>(.167)<br>.156 | .292<br>(.166)<br>.080 |  | .209<br>(.196)<br>.288 | .238<br>(.193)<br>.218 | .305<br>(.191)<br>.111 |
| Anxiety-Depression-Stress<br>Scale |  | .082<br>(.021)<br><.001 | .081<br>(.021)<br><.001 | .091<br>(.021)<br><.001 |  | .230<br>(.023)<br><.001 | .281<br>(.023)<br><.001 | .217<br>(.023)<br><.001 |  | .189<br>(.023)<br><.001 | .182<br>(.022)<br><.001 | .191<br>(.022)<br><.001 |
| Female |  |  | .784<br>(.136)<br><.001 | .879<br>(.135)<br><.001 |  |  | .671<br>(.134)<br><.001 | .762<br>(.135)<br><.001 |  |  | .867<br>(.148)<br><.001 | .968<br>(.147)<br><.001 |
| Non-White |  |  | .574<br>(.142)<br><.001 | .652<br>(.146)<br><.001 |  |  | .528<br>(.141)<br><.001 | .505<br>(.145)<br><.001 |  |  | .232<br>(.154)<br>.133 | .244<br>(.158)<br>.124 |
| Post-Secondary<br>Degree |  |  | 1.215<br>(.137)<br><.001 | .927<br>(.142)<br><.001 |  |  | 1.165<br>(.134)<br><.001 | .960<br>(.139)<br><.001 |  |  | 1.362<br>(.148)<br><.001 | 1.060<br>(.153)<br><.001 |
| Urban Location |  |  |  | .409<br>(.156)<br>.009 |  |  |  | .439<br>(.153)<br>.004 |  |  |  | .613<br>(.167)<br><.001 |
| Household income<br>greater than<br>\$50,000 annually | | | | .860<br>(.147)<br><.001 | | | | .615<br>(.142)<br><.001 | | | | .902<br>(.156)<br><.001 |
| Household member<br>with COVID-19<br>diagnosis |  |  |  | .897<br>(.690)<br>.193 |  |  |  | .867<br>(.616)<br>.159 |  |  |  | -1.528<br>(.556)<br>.006 |
| Adjusted R-squared | .009 | .012 | .064 | .081 | .004 | .046 | .089 | .101 | .004 | .036 | .090 | .113 |

**Notes:** Cell entries are regression coefficient B, (SE), and P-value.
**Mitigation compliance scale** summed 17 items: washing or sanitizing hands, wiping packages entering the home, wearing face masks, staying at home when unwell, keeping distance from individuals outside the household, postponing or cancelling doctor appointments, dental appointments, housekeeper or caregiver services, recreational activities, work activities, school activities, avoiding going to restaurants, public or crowded places, avoiding contact with high-risk people, stockpiling of food or water, working from home, and studying from home. Scale reliability was 0.72 – 0.80.
**Anxiety-depression-stress scale** summed days of feelings of depression, anxiety, hopelessness, and loneliness, and stress experienced by CIS respondents within the last 7 days. Scale reliability was 0.81 – 0.86.
**Symptoms score** summed fever, chills, shortness of breath (dyspnea), cough, sore throat, fatigue or tiredness, muscle pain (myalgia), and loss of taste and smell (anosmia).
**Risk factors score** summed diabetes, high blood pressure or hypertension, heart disease or stroke, asthma, chronic lung disease or COPD, bronchitis or emphysema, liver disease, cancer, and immunocompromised condition.

## Discussion

The pandemic prompted strong mitigation behavior by adults, especially among females, non-whites, urban dwellers, and the psychological unwell. Other positive predictors were post-secondary education and higher income. Health symptoms and clinical risk factors did not predict increased mitigation practices, nor did age 65+ and proximity to infected persons. Our study findings are congruent with a report that pointed to a lack of increased pandemic mitigation practices in households with confirmed infections and health risks.^1^ Our results also point to lower levels of psychological resilience, lower socio-economic status, and non-urban location as potential explanatory factors for lack of mitigation behavior.

Limited data were available for our study and further research is needed to explain mitigation behavior. The CIS may have sampling bias and the questionnaire did not support the computation of previously validated psychological scores. The anxiety-depression-stress scale may only be suitable for this epidemiologic study. Lastly, we only validated scale reliability using Chronbach’s alpha.

## Data Availability

Our study used public COVID Impact Survey data.

https://www.covid-impact.org/

